# Remote working and experiential wellbeing: A latent lifestyle perspective using UK Time Use Survey before and during COVID-19

**DOI:** 10.1101/2022.04.27.22273297

**Authors:** Jerry Chen, Li Wan

**Author notes:** Corresponding author: Dr Li Wan, Corresponding.

## Abstract

Mental health in the UK had deteriorated compared with pre-pandemic trends. Existing studies on heterogenous wellbeing impacts of COVID-19 tend to segment population based on isolated socio-economic and demographic indicators, typically gender, income and ethnicity, while a more holistic understanding of such heterogeneity among the workforce seems lacking. This study addresses this gap by 1) combining UK time use surveys collected before and during COVID-19, 2) identifying nuanced lifestyles within three working mode groups (commuters, homeworkers and hybrid workers) using latent class model, and 3) quantifying heterogeneous experiential wellbeing (ExWB) impacts across workers of distinct lifestyles. It was found that the direction and magnitude of ExWB impact were not uniform across activity types, time of day and lifestyles. ExWB impact outside of usual working hours (before 6am and after 6pm) was consistently negative for all lifestyles. In contrast, the direction of ExWB impact during usual working hours (6am-6pm) varied in accordance with lifestyle classifications. Specifically, ExWB decreased for all homeworkers but increased significantly for certain hybrid workers. Magnitude of ExWB impact correlated strongly with lifestyle: the optionality of choosing one’s place of work and the associated ability to travel during the pandemic seemed to improve ExWB. To understand the significant heterogeneity in ExWB outcomes, a spatial-temporal conceptualisation of working flexibility is developed to explicate the strong yet complex correlations between wellbeing and lifestyles. Whilst greater spatio-temporal flexibility is generally linked to increase in workers’ ExWB, there is preliminary evidence of a flexibility threshold – above which the marginal ExWB increase would diminish and even become negative. The implications to post-pandemic “back-to-work” policies are 1) shifting policy focus from simplistic workplace choice to spatial-temporal optionality (i.e. lifestyle choice), and 2) providing wider support for lifestyle adaptation and transitions.

## INTRODUCTION

Mental health in the UK had deteriorated compared with pre-pandemic trends (Pierce et al, 2021). Early impact of COVID-19 on subjective wellbeing was primarily caused by increased psychological distress from emergency lockdown measures (Niedzweidz et al., 2021; Holmes et al., 2021). While post-pandemic recovery progresses across the globe, prolonged social distancing and travel restrictions and social scarring have led to profound societal changes (Batty, Clifton, Tyler, & Wan, 2022; Florida, Rodríguez-Pose, & Storper, 2021) – particularly concerning the future of work (Peters et al., 2022). In the UK, rapid democratisation of remote working has been instrumental in maintaining productivity of firms during the pandemic and flexible/remote working is likely to remain in practice for some occupations and sectors (UK Parliament, 2022). Home-based and hybrid working is not new; however, the wellbeing impact of flexible/remote working is not fully understood yet further complicated by our lived experience of the pandemic. Existing studies on heterogenous wellbeing impacts of COVID-19 tend to segment population based on single socio-economic and demographic indicators, notably gender (Tan & Lim-Soh, 2023), income (Martinez et al., 2020) and ethnicity (Zhou & Kan, 2021), while a more holistic, evolving understanding of such heterogeneity among the workforce seems lacking.

Time-use data provides a unique source of information for understanding nuanced and evolving wellbeing impacts of COVID-19 (Blanchflower, 2008; National Research Council, 2013; OECD, 2013; Lee & Tipoe, 2021). This paper aims to address the above research gap by empirically identifying heterogeneous wellbeing impact of flexible/remote working across three dimensions, 1) aggregate daily activity types, 2) latent lifestyles of workers and 3) the evolution from pre-to during the pandemic. While the first dimension on activity types has been explored in the literature (Lee & Tipoe, 2021), the lifestyle perspective and the longitudinal dimension combined are new thus contribute to our understanding of the impact of flexible/remote working on experiential wellbeing. In terms of analytical strategy, latent class model is applied to identify nuanced lifestyles within each of the three working mode groups (homeworkers, commuters and hybrid workers). The novel use of latent class model on time-use data allows efficient and holistic segmentation of workers based on their socio-economic and demographic profiles and daily activity patterns. The longitudinal dimension is enabled by combining population-representative, repeated cross-sectional time use data gathered during the pandemic (2020) with the pre-pandemic UK Time Use Survey (UKTUS) in 2015.

Based on findings from the lifestyle perspective, we further propose a two-dimensional framework for conceptualising ‘working flexibility’ in an intraday setting, which helps explicate strong yet complex correlations between wellbeing and daily activity pattern. The new framework builds on the work of Antilla et al. (2015), featuring a separation of spatial and temporal dimension for measuring intraday working flexibility based on time-use data. *Temporal flexibility is often overlooked as spatial flexibility dominates existing discourse on flexible working practices*. The incorporation of the temporal dimension in conjunction with the lifestyle perspective is expected to shed light on the relationship between spatial-temporal working arrangements and subjective wellbeing. Implications for ‘back-to-work’ policies are discussed, emphasising the policy need for facilitating lifestyle transitions, as opposed to simplistically reverting the location of work.

## LITERATURE REVIEW

### Measurements of subjective wellbeing

Leveraging the strength of the time-use data, subjective wellbeing is conceptualised as experienced or experiential wellbeing (ExWB) (Stone et al., 2018). ExWB, sometimes referred to as hedonic wellbeing, is the series of momentary affective states that occur at one or a series of point-in-time reference periods (National Research Council, 2013). Typical questions measuring ExWB are “How do you feel at this moment?” or “How did you enjoy commuting to work yesterday morning?”. In contrast with evaluative wellbeing, which involves comparatively long periods and focuses on global assessments on one’s state of being or satisfaction through e.g. the 12-item General Health Questionnaire (GHQ-12), ExWB measurement aims to capture a respondent’s immediate focus, rather than broader issues that fall in the domain of evaluative wellbeing (National Research Council, 2013).

Compared to evaluative wellbeing, ExWB has been the lesser-studied aspect of subjective wellbeing. The reasons are twofold. Firstly, the point-in-time reference period likens ExWB to a fleeting emotional state, thus have questioned the volatility and clinical relevance of ExWB (Kahneman, 2004). However, the temporal aspect is a unique strength of ExWB, as it captures emotions as they fluctuate from moment to moment and in response to day-to-day events and activities (OECD, 2015). Secondly, despite the ability to capture instantaneous wellbeing, collecting ExWB data is resource intensive (Kreuger et al., 2009). For the measurement to have explanatory power, measurements associated with different time points and different activities throughout time need to be gathered. This requires a herculean effort of data collection and poses a heavy burden for the respondents as well (Cornwall et al., 2019). Our research takes advantage of the high-granularity and population-representative ExWB data in the UKTUS data. The wealth of ExWB data available both pre and during the pandemic is a novel enabler for our research.

### Why are we focusing on time use?

During the pandemic, significant changes in both quantity and quality of time spent on different activities have been observed. Lee & Tipoe (2021) found that individuals tended to spend more time on housework and less time on paid work activities during COVID-19. They also found that in the third lockdown, one fifth of the samples experienced increased atypical working hours, which strongly correlates with decreases in their ExWB. The disutility associated with increased atypical paid work hours is compounded by unenjoyable substitution activities taken place during typical hours, such as non-paid work (e.g. childcare) or alone leisure time. Lee & Tipoe (2021) called for further research on the factors that affects ExWB beyond the specific activity conducted.

Adam et al. (2018) leveraged the UKTUS 2015 data to study experiential wellbeing levels during commuting episodes. Averaging across all samples, time spent in passive commuting and paid work are associated with the lowest level of subjective wellbeing. The authors compared commuting days to non-commuting days, and found that after controlling for usual working arrangement and other socioeconomic characteristics, non-commuting days tend to lead to higher reported ExWB levels. The findings are consistent with an earlier study by the ONS (2014). However, the authors found that notably there were no meaningful differences in ExWB levels for paid work activities between commuting and non-commuting days prior to the pandemic.

Zhou & Kan (2021) analysed subjective wellbeing levels during three lockdown periods and the intermedial easing periods. Subjective wellbeing is measured through the GHQ-12 from the UK Household Longitudinal Survey COVID studies. The authors focused on the change in distress levels throughout the one-year period between April 2020 and March 2021. While time use patterns became less sensitive to the later lockdowns, distress levels reached a new high after repeated lockdowns. These negative wellbeing impacts are heterogeneous across social groups. Subjective wellbeing decreased more rapidly for women, whereas men experienced the deterioration in wellbeing more gradually, but reached higher levels of distress progressively with each subsequent lockdown. Women recovered their wellbeing faster, and maintained a similar peak level of distress in all three lockdowns. BAME (Black, Asian, Middle Eastern) minority groups experienced greater distress in the first lockdown.

### An evolving definition of flexible working

It is well researched that flexible working practices such as remote working enhances the sense of autonomy, job satisfaction, and other evaluative wellbeing measures (Anderson et al., 2015; Fonner & Roloff, 2010; Gajendran & Harrison, 2007). However, COVID-19 profoundly changed the definition of flexible working. Previous studies on the COVID-19 impact on workers’ wellbeing are limited to specific activities (e.g. paid work in Barrero, Bloom & Davis, 2021), or specific demographic groups (e.g. young adults in Gagne, Nandi & Schoon, 2021). Few studies explicitly compared ExWB changes of distinct worker groups such as commuters, homeworkers, and hybrid workers. Cross-sectional data during the first lockdown in Portugal showed that homeworkers had a higher level of job satisfaction, due to better work-life balance and flexibility (Sousa-Uva et al., 2021). Among homeworkers there were yet more distinct groups with varied wellbeing outcomes. A multi-national European study showed occasional homeworkers had high levels of satisfaction with their job quality, while highly mobile homeworkers reported poor work-life balance (Rodriguez-Modroño & Lopez-Igual, 2021). There was also evidence suggesting heterogeneous pandemic effects on mental health based on work environments (Ervasti et al, 2021).

A systematic review of ten controlled before and after studies found that increasing worker control and choice through flexible work interventions were likely to have a positive effect on health outcomes, but there was a clear need to delineate the impact of flexible working on wellbeing (Joyce et al., 2010). National lockdowns in the UK were widely imposed across demographic and socioeconomic groups. However, studies involving occupational cohorts have largely been restricted to healthcare and/or essential workers (Carr et al., 2021). There is a clear gap in literature for a study that looks at the entire working population, to compare across population subgroups with different working arrangements, and to account for latent yet distinct lifestyles within these subgroups. Lifestyle differences and the shift to more flexible working practices enabled by remote work need to be accounted for to delineate the impact of COVID-19 on ExWB. Through combining the pre- and during pandemic Time Use Survey data, this study aims to quantify the changing relationship between time use and wellbeing across heterogenous yet latent groups of employed workers in the UK.

## DATA AND METHODS

### Data source

UKTUS (2015) is a large-scale household survey that provides data on how people spend their time. Time diaries record activity sequences, corresponding locations, as well as the respondent’s level of *enjoyment* for every activity episode recorded throughout the day (Gershuny & Sullivan, 2016). We combined the pre-pandemic UKTUS 2015 with four additional waves of population-representative time use data collected during the pandemic, which is novel in time-use literature. These waves were collected online in 1) 2016; 2) May-June 2020 at the peak of first UK COVID-19 lockdown; 3) August 2020 following the relaxation of social restrictions; and 4) November 2020 during the second lockdown (Gershuny & Sullivan, 2021). The latter three waves collected throughout 2020 were classified in our analysis as ‘during COVID’, while the ‘pre-COVID’ data consisted of UKTUS 2015 and the first online wave collected in 2016. All data was obtained from the UK Data Service.

The study samples comprised all those aged 16-64, in full employment, with sufficient data on activity-level episodic ExWB, and a non-zero survey weight. Our analysis leverages the unique strength of UKTUS 2015 and its 4-wave extensions during COVID-19, which include 3,855 individuals, each contributing 24 hours of time use schedule during a typical weekday in 10-minute episodes. A total of 369,394 activity-location-ExWB bundles were analysed (see sample distribution in Appendix 1).

### Measures

Prior to endogenous identification of lifestyles using the latent class mode, we first separated all samples by the self-reported location of paid work into three distinct working modes: 1) homeworkers; 2) commuters; and 3) hybrid workers. Homeworkers engaged in paid work exclusively at home during the diary day; commuters engaged in paid work exclusively at the workplace; whilst hybrid workers reported paid work activities both at home and in the workplace. Such information had been collected consistently across survey periods. Other sociodemographic characteristics considered were income, educational attainment, occupation classification, gender, age, and marital status.

UKTUS has 144 distinct activity categories. To reduce the complexity, we followed literature (Lee & Tipoe, 2021) and aggregated them into broad activity types: 1) personal; 2) paid work; 3) non-paid work; 4) leisure; and with our addition of 5) transport (see breakdown and aggregation methods in Appendix 2). Intraday time use pattern was also aggregated from 10-minute intervals into 24 hourly variables, based on the broad activity type a respondent was engaged in for the majority of a given hour.

A unique strength of UKTUS is the granular experiential wellbeing (ExWB) measurement based on the *enjoyment* variable. We recognise the limitation of using a single-measure subjective wellbeing, but the selection of subjective wellbeing measures represents a difficult trade-off. While a multi-dimensional wellbeing measure (e.g. GHQ-12) would be more comprehensive and comparable across countries, the *enjoyment* measurement in UKTUS provides a high level of granularity that captures the intraday fluctuations of wellbeing, which is unique and worth further investigation. Arguably, the experiential wellbeing data in UKTUS seems to have been underutilised in literature given its uniqueness and granularity.

For every activity, a respondent’s ExWB on the 7-point Linkert scale is obtained, 1 represents “Did not enjoy at all”, 2 represents “Did not enjoy”, 3 represents “Slightly did not enjoy”, 4 represents “Neutral”, 5-7 mirrors the ordinal pattern for positive responses. Our descriptive analysis found that the distribution of ordinal responses was heavily right skewed, where the proportion of the top two scores (i.e. 6 – “Enjoyed” and 7 – “Enjoyed very much”) accounting for roughly half of overall responses (see distribution histogram in Appendix 3). The significant skewness of the ExWB measure, if not corrected, would lead to biased model estimations. In this paper, the ExWB measure was transformed into a binary variable with *enjoyment* six and above defined as high ExWB, five and below defined as average-to-low ExWB. The use of binary ExWB measurement seems novel in literature and has two advantages. Firstly, the ordinal enjoyment measurement is difficult to compare across years and individuals, given its highly context-dependent nature. Using a binary variable could, to some extent, reduce the volatility and increase the clinical relevance of enjoyment measurement by considering the relative level of ExWB. It enabled us to focus on identifying determinants that may lead to higher-than-average ExWB, as opposed to factors that may lead to a general increase of enjoyment. Secondly, it reduces the computational complexity for model estimation.

### Statistical analysis

*Latent class analysis (LCA)* is used to identify nuanced lifestyles within each working mode group (homeworkers, commuters and hybrid workers). We estimated latent class models using robust maximum likelihood estimation with Newton-Raphson stepping mechanism, repeated for one to six latent classes. The 24 hourly categorical variables indicating activity type were used as explanatory variables in the models, together with socioeconomic covariates including income quantile, education attainment, and occupation classification. To help with model convergence, we used a two-step integration process: Laplacian approximation was used in the first round of integration, the results matrix was then used as starting value for the more accurate mean-variance adaptive Gauss-Hermite quadrature integration method. The final class size within each working mode was determined using the Akaike’s information criterion and Bayesian information criterion values (see model fit estimations in Appendix 4), average latent class probabilities, and substantive interpretation of the classes identified. The LCA was conducted using pooled samples from both pre- and during COVID-19. After LCA, the samples were then separated into pre- and during COVID groups according to the survey date.

*Logistical regression models* are used to explore potential factors that contribute to higher-than-average ExWB. The dependent variable is the binary ExWB (1 for higher ExWB and 0 for lower and average ExWB), and the independent variables in the model are broad activity type, lifestyle, the interaction term between broad activity type and lifestyle; socioeconomic covariates include sex, age group, income group, education attainment, occupation classification and residential region. Month of survey collection is added to control for idiosyncratic differences. Results for pre- and during COVID data are compared.

### Conceptualising spatio-temporal flexibility

A two-dimensional conceptual framework for measuring working flexibility is proposed, aiming to explicate the heterogenous wellbeing impacts across different lifestyles of workers. The spatial dimension measures the worker’s ability to choose the location to engage in paid work on an intraday basis. *Temporal flexibility* is a distinct but interrelated dimension of flexibility, which concerns the ability to choose when to conduct paid work on an intraday basis, which in turn impacts how they organise their non-work activities including travel.

The temporal flexibility is quantified using a single entropy measure of when breaks related to Personal and Leisure activities take place during the working hours, e.g. lunch breaks and short strolls. The daily working hours is identified by the timestamp of the first and last episode of paid work. One will achieve high temporal flexibility if they engage in numerous or prolonged breaks during the working hours.

## RESULTS

A total of ten lifestyles were identified across the three working modes: three among homeworkers, four among commuters and three among hybrid workers. Following the tempogram method developed by Kolpashnikova et al., (2021), Figure 1 shows the distinct intraday time use patterns of different lifestyles and the corresponding intraday ExWB patterns. Table 1 further provides the pen portrait for each lifestyle. The naming of *lifestyles* is primarily based on paid work schedules because of its dominant role in lifestyle choice and identity (Antilla et al., 2015). Sociodemographic characteristics of each lifestyle are summarised in Appendix 5.

**Figure 1:**
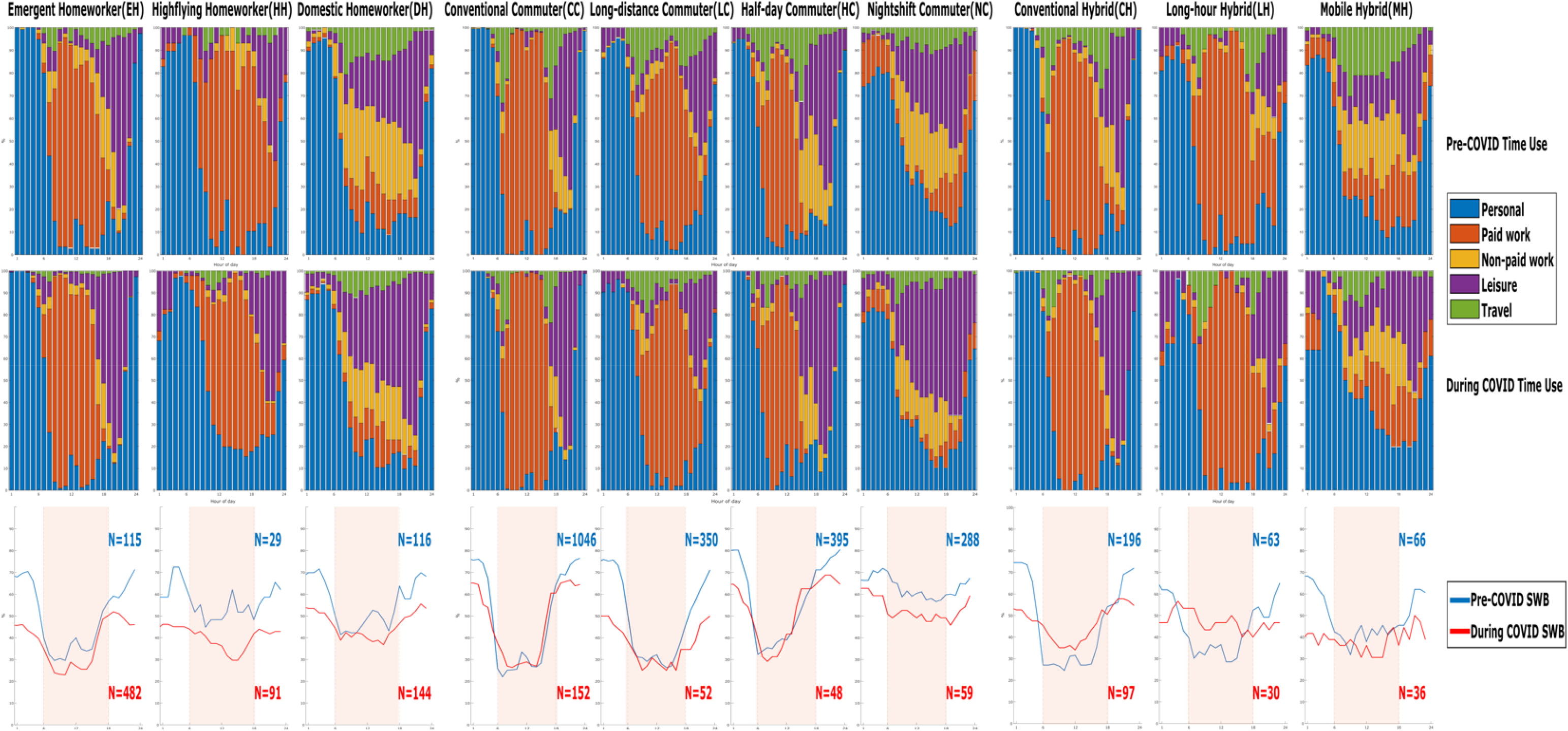
Intraday patterns of time use (top two rows) and ExWB (bottom row) *

**Table 1:** Lifestyle pen portraits

| Working Mode |  | Homeworker | Commuting Worker | Hybrid Worker |
| --- | --- | --- | --- | --- |
|  |  | Paid work engaged exclusively at home. Younger and single demographics increased during COVID-19. | Paid work engaged exclusively at workplace. Demography became older and generally worse off economically during COVID-19, most of whom are essential workers. | Paid work split between home and workplace. The proportion of respondents in hybrid work mode remained consistent pre- and during COVID-19. |
| Lifestyle | Early Concentrated Working | ***Emergent Homeworker (EH)***<br>Emerging and dominant lifestyle during COVID-19 representing 40% of all respondents (70% of Homeworkers), a tenfold increase from its pre-pandemic levels. | ***Conventional Commuter (CC)***<br>Dominant lifestyle pre-pandemic representing 39% of all respondents (50% of Commuters). | ***Conventional Hybrid (CH)***<br>Highly male dominated both pre- and during COVID-19; dominant lifestyle within the hybrid work mode (60%). |
|  | Long Dispersed Working | ***Highflying Homeworker (HH)***<br>Highest share of high-income earners; little time spent on commuting and more time on paid work and personal maintenance. | ***Long-distance Commuter (LC)***<br>Most time spent on commuting, and very little time on non-paid work; younger, with leisure activities last late into the evening. | ***Conventional Hybrid (CH)***<br>Highly male dominated both pre- and during COVID-19; dominant lifestyle within the hybrid work mode (60%). |
|  | Non-work Dominated | ***Domestic Homeworker (DH)***<br>Least amount of time spent in paid work and most time spent in non-paid work both pre- and during COVID-19. One of the few lifestyles dominated by female. |  | ***Long-hour Hybrid (LH)***<br>The only lifestyle spending more than 10 hours in paid work both pre- and during COVID-19; little time for sleep and other personal maintenance. |
|  | Half-day Working |  | ***Half-day Commuter (HC)***<br>High proportion of manual workers, and low proportion of managers; lowest proportion of undergrad degree holders; demography was heavily skewed toward older age groups. |  |
|  | Nightshift |  | ***Nightshift Commuter (NC)***<br>Greatest decrease of high earners during COVID-19, as a result, it has the highest portion of low earners. |  |

The defining time use pattern for each lifestyle remained largely consistent pre- and during COVID-19, though the sample distribution across lifestyles changed significantly, notably a significant shift from Conventional Commuter (CC, N=1,046 pre-COVID) to Emergent Homeworker (EH, N=482 during COVID). The direction and magnitude of pandemic impact on ExWB were heterogenous across time of the day, activity types and lifestyles. We discuss these aspects in turn.

### Time use and ExWB patterns outside usual working hours (before 6am or after 6pm)

ExWB declined uniformly across lifestyles outside usual working hours when most people engaged in leisure or personal activities such as sleeping. However, time use pattern during these hours remained virtually unchanged pre- and during COVID, which suggests that the ExWB decline during these hours seemed uncorrelated with time use and may be attributed to general anxiety and potentially blurred boundary between work and life associated with flexible working. This finding emphasises the need to investigate nuanced intraday variation of subjective wellbeing through separating usual working hours from the rest of the day. It could be postulated that the decline of ExWB outside usual working hours was a main contributor to the overall deterioration of mental health in the UK during COVID-19. Furthermore, a rapid and significant rebound of ExWB after work (around 6pm) was observed consistently across lifestyles before the pandemic. During COVID-19, however, the rate of ExWB rebound in the early evening flattened, particularly for homeworkers and hybrid workers who tend to be more susceptible to the effect of blurred boundary between work and life.

### Time use and ExWB patterns within usual working hours (6am-6pm)

The negative ExWB impact of paid work activities were reflected in the majority of lifestyles. Enjoyment tended to plummet as usual working hour started, with slight uptick in mid-day which coincided with usual lunch breaks, and some recovery after work. The uptick during lunch break was notably less notable during COVID-19 and even disappeared for some lifestyles (e.g. HH, DH and CH). Lifestyles with more concentrated paid work periods such as EH, CC, and CH would experience greater intraday ExWB fluctuations. To quantify the varying impacts while controlling for sociodemographic covariates, logistical regression is applied to activity episodes within the usual working hour window. Results are reported in Table 2.

**Table 2:** Logistical regression models – Pre-vs During COVID-19 (within usual working hours & workdays only)

|  |  | **Pre-COVID-19** |  | **During COVID-19** |  |
| --- | --- | --- | --- | --- | --- |
|  |  | Coefficient | Robust standard error | Coefficient | Robust standard error |
| **Broad Activity Type** | Personal | **0.54\*\*\*** | 0.04 | **0.98\*\*\*** | 0.06 |
|  | Paid Work | **Ref** | - | **Ref** | - |
|  | Non-paid Work | **0.34\*\*\*** | 0.05 | **1.03\*\*\*** | 0.10 |
|  | Leisure | **1.23\*\*\*** | 0.07 | **1.34\*\*\*** | 0.09 |
|  | Transport | **-0.05** | 0.05 | **0.55\*\*\*** | 0.07 |
| **Lifestyle** | EH | **0.21\*\*** | 0.09 | **-0.15\*\*\*** | 0.03 |
|  | HH | **0.84\*\*\*** | 0.14 | **0.12\*\*\*** | 0.05 |
|  | DH | **0.26\*\*** | 0.13 | **-0.12** | 0.07 |
|  | CC | **Ref** | - | **Ref** | - |
|  | LC | **-0.24\*\*\*** | 0.06 | **-0.17\*\*\*** | 0.06 |
|  | HC | **0.33\*\*\*** | 0.06 | **0.19\*\*\*** | 0.06 |
|  | NC | **0.22\*** | 0.13 | **-0.37\*\*** | 0.18 |
|  | CH | **-0.09** | 0.06 | **0.34\*\*\*** | 0.04 |
|  | LH | **0.16\*** | 0.10 | **0.39\*\*\*** | 0.06 |
|  | MH | **0.15** | 0.17 | **-0.39\*\*\*** | 0.12 |

\* $p < 0.05$ , \*\* $p < 0.01$ , \*\*\* $p < 0.001$

Before the COVID-19 pandemic, homeworkers, particularly the highflying homeworkers (HH), tended to have better ExWB than conventional commuters (CC, as the reference case). ExWB of hybrid workers was not statistically higher than CC. It implies that homeworking may be a privilege for some (e.g. HH), but hybrid working appeared a compromise for those who had to juggle paid work and domestic duties. However, during COVID-19, relative ExWB (based on CC) of all homeworker groups decreased significantly, particularly the EH group, the majority of whom were likely new homeworkers transitioned from commuters as a result of lockdown measures. By contrast, relative ExWB increased for all hybrid lifestyles except the Mobile Hybrid (MH), who experienced a significant decline of ExWB relative to CC. MH lifestyle features the shortest paid work and longest non-paid work both pre- and during COVID-19 (see Appendix 5). The decrease of ExWB for homeworkers and increase of ExWB for hybrid workers combined suggest that the optionality over place of work and the ability to continue to travel during the pandemic seemed to bring ExWB betterment. However, for hybrid workers who bear considerable domestic duties, the positive ExWB effect associated with such optionalty would diminish and may even turn into negative if fulfilling certain domestic duties (e.g. home schooling, caring duties outside own home) became increasingly difficult during the pandemic.

Other plausible factors that may contribute to the ExWB increase of hybrid workers during the pandemic include social interaction opportunities at the workplace, the ability to stay outdoor as part of travel and the formalisation of hybrid working as an acceptable practice. *It is also found that the ExWB increase for hybrid workers stem from enjoyment boost associated with some activity types but not all, which will be investigated in the next section*.

### Heterogeneity of ExWB impact across activity types and latent lifestyles

Figure 2 shows average ExWB changes from pre-to during COVID-19 within usual working hours (6am-6pm) by activity type and lifestyle. In terms of the direction of ExWB impact, ExWB for paid work decreased for almost all lifestyles, with a notable exception of the long-hour hybrid lifestyle (LH), which might be attributed to the increasing appreciation of certain occupations during the pandemic. ExWB decline on leisure activities was observed for almost all lifestyles, despite an increase of time spent for leisure for all lifestyles (see Appendix 5). Lee & Tipoe (2021) pointed out that the lack of company and social interactions in leisure activities may be a main cause of ExWB decline.

**Figure 2:**
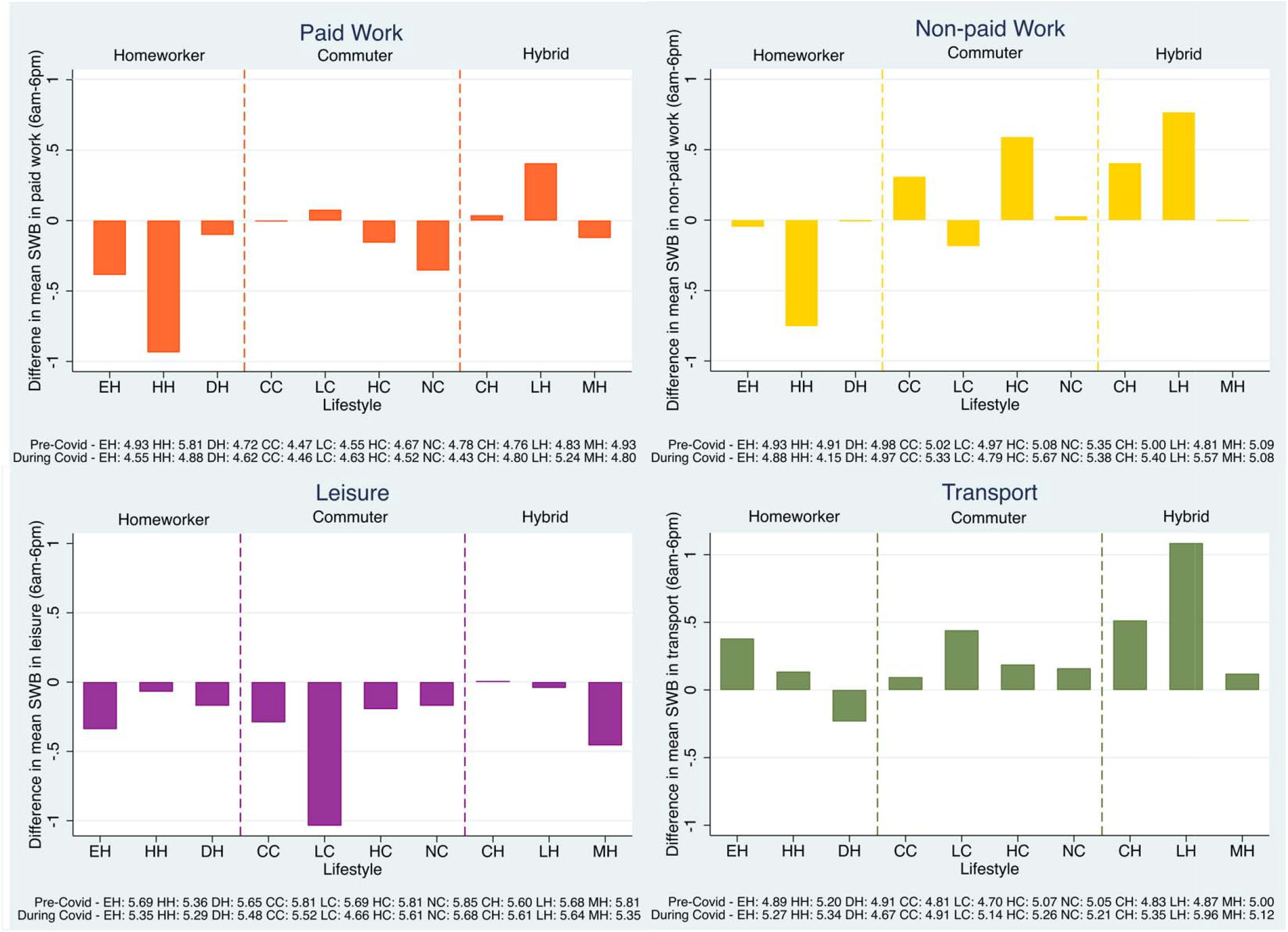
Difference in mean ExWB within usual working hours (6am-6pm) by activity type * The y-axis represents the difference in average ExWB for each lifestyle while engaged in the respective

By contrast, ExWB associated with travel activities has increased for most lifestyle groups except the Domestic Homeworker (DH), who spent more time on transport than other homeworkers and on par with some commuters (see Appendix 5). For DH, the need to engage in essential trips (e.g. to pharmacies and caring duties) may cause stress and anxiety due to difficulties in travel and the risk of contagion. ExWB increase from moderate travel activities, particularly for hybrid workers (CH & LH) during the pandemic corroborates the early finding that the optionality over place of work and the ability to continue to travel had a positive ExWB effect. For non-paid work, ExWB increased for some commuter and hybrid worker lifestyles, but decreased for all homeworker groups, implying emerging difficulties in (re-)balancing life and work for homeworkers.

In terms of the magnitude of COVID-19 impact, five out of six large ExWB changes (>= 0.5 or <=-0.5) occurred in late and dispersed lifestyles: HH, LC, and LH. This is contrasted by the relatively low magnitude of impact for the non-work dominated lifestyles: DH, HC, NC, and MH. These lifestyles tended to be less constrained by established work-time regime, enabling a higher degree of flexibility for adjusting their time use to mitigate the pandemic impact. activity type. Pre- and during COVID-19 average values used to calculate the differences are in the lege d.

### Conceptualising working flexibility – A time use perspective

Our analysis shows COVID-19 impact on ExWB is heterogeneous in terms of the direction and magnitude for different activity types and lifestyles. The identification of latent lifestyles and the comparison of ExWB impact across lifestyles shed a new light on the conceptualisation of ‘flexibility’ from a time use perspective. Specifically, *flexibility could be conceptualised over two dimensions, spatial flexibility and temporal flexibility* (Antilla et al., 2015). *Based on two normalised entropy measures for each reflecting spatial and temporal flexibilities, we visualise a range of spatio-temporal flexibilities for each lifestyle in Figure 3*.

**Figure 3:**
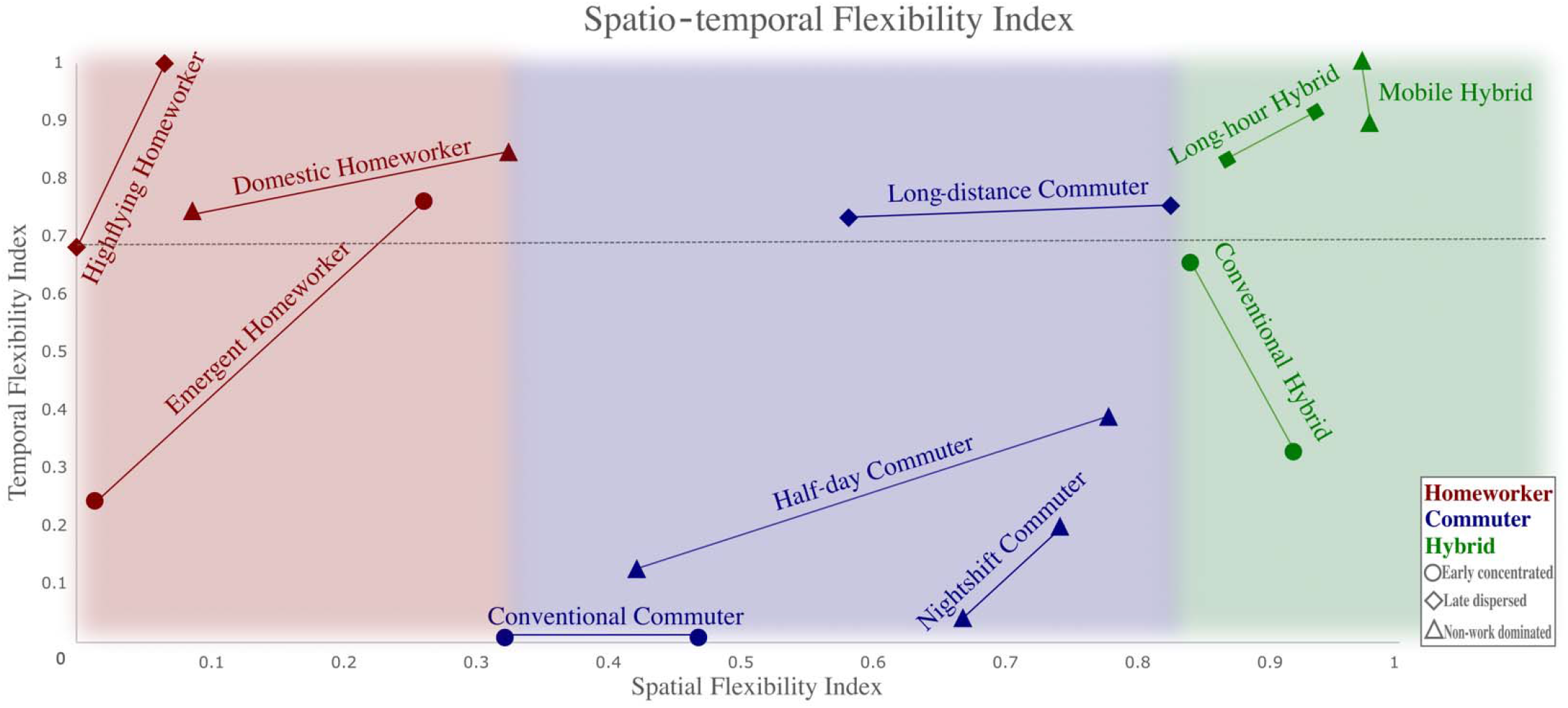
Spatio-temporal flexibility by working arrangement & lifestyle (adopted from Wan & Chen, 2022) * The axes are normalised entropy measures for spatio-temporal flexibilities. Each lifestyle has two data points, one calculated using data from pre-COVID-19 data and the other during COVID-19. The two points are connected to represent the range of spatio-temporal flexibility between pre-pandemic normalcy and changes induced by COVID-19.

Spatial flexibility is a normalised entropy measure of where workers are located during the working hours. The key variable measured is the location of an individual between the first and last hour for paid work. All locations recorded within the period are included in the entropy calculation, regardless of activity type. Spatial flexibility is closely associated with working modes – homeworkers (hybrid workers) tend to have the lowest (highest) spatial flexibility as depicted along the horizontal axis in Figure 3.

Temporal flexibility is a normalised entropy measure of when potentially enjoyable breaks take place over the working day. Potentially enjoyable breaks are defined as Personal or Leisure activities during the working day (e.g. lunch break or a short stroll). A higher entropy would derive from numerous or prolonged enjoyable breaks, indicating greater ability to organise one’s time, hence greater temporal flexibility. Temporal flexibility is closely associated with lifestyle – all three long and dispersed working lifestyles (HH, LC, LH) and two non-work dominated lifestyles (DH & MH) have consistently high temporal flexibility pre- and during COVID-19.

The spatio-temporal framework provides a novel perspective to understand the potential wellbeing impact of flexible working. Existing discourse on flexible working has been predominantly focused on the spatial dimension (e.g. in the form of hybrid working), with an underlying assumption that the higher flexibility the better for workers. However, our analysis suggests that hyper spatial flexibility, combined with hyper temporal flexibility, may be detrimental to ExWB. For example, the Mobile Hybrid (MH) lifestyle featured high flexibility in both dimensions, but their ExWB was not statistically higher than Conventional Commuters (CC) pre-pandemic (see Table 2). During the pandemic, the MH lifestyle (mostly married, young respondents – see Appendix 5) experienced a notable decrease in ExWB relative to CC. By contrast, lifestyles of relatively high temporal flexibility and low-to-moderate spatial flexibility showed positive ExWB effect pre-COVID-19.

The implication is twofold: firstly, the marginal utility of greater spatio-temporal flexibility diminishes and may even turn to negative (e.g. working across many locations to the point that breaks are no longer enjoyable but a mere necessity); secondly, the hyper spatial-temporal flexibility, as captured by the proposed measurements, may represent a compromise, rather than a voluntary lifestyle choice. The negative wellbeing impact of such involuntariness exacerbated during the pandemic, potentially due to imposed furlough and/or increasing domestic duties such as home-schooling.

To postulate wellbeing impact of flexible working in a post-pandemic context, spatial flexibility is likely to increase for all lifestyles due to recovered confidence for travel, while the change of temporal flexibility tends to be more complex across lifestyles. On the one hand, relatively high temporal flexibility as an outcome of voluntary lifestyle choice may explain the positive wellbeing effect, hence the desirability of homeworking and hybrid working, but such positive effect may be rather limited if a more temporally flexible working pattern was not offered by employers. On the other hand, the non-linear relationship between spatial-temporal flexibility and ExWB suggests that workers may benefit from the optionality of multiple working modes across days of the week, e.g. reduced (increased) working hours for some (other) days of the week. The lifestyle perspective based on time-use patterns seems effective to capture such nuances.

### Conclusion

This paper aims to investigate nuanced and evolving wellbeing impacts of COVID-19 across distinct lifestyle groups of workers in the UK. The main strength of the study is our novel lifestyle perspective enabled by the time-use survey data collected before and during COVID-19. Through identifying latent but distinct lifestyles of workers in the UK, our research provides timely and fresh insights on the wellbeing impact of flexible working and the heterogenous impact of COVID-19 across activity types and lifestyles. It was found that the direction and magnitude of ExWB impact were not uniform across activity types, time of day and lifestyles. ExWB impact outside of usual working hours (before 6am and after 6pm) was consistently negative for all lifestyles. In contrast, the direction of wellbeing impact during usual working hours (6am-6pm) varied across lifestyles. It is also demonstrated that a spatial-temporal conceptualisation of working flexibility may help explicate the strong and non-linear correlations between ExWB and lifestyles. While flexible working (e.g. in the form of 4-day work week) is gaining and retaining popularity for some sectors/occupations (Whillans & Lockhart, 2021), our empirical finding contests the simplistic assumption that the higher working flexibility the better. Homeworking is not inherently more flexible, especially when temporally inflexible work-time regime persists. The non-linear relationship identified emphasises the imperative of developing a proper conceptualisation of working flexibility, which would be crucial for informing the ‘back-to-work’ policies.

One key policy implication is that existing discussion on ‘back-to-work’ policies should expand beyond the spatial dimension of working flexibility (i.e. where to work) and focus more on the temporal dimension in relation to more nuanced segmentations of the workforce. Interventions to increase temporal working flexibility may lead to higher worker wellbeing and resilience to tackle negative shocks in the labour market. A lifestyle perspective based on time-use patterns seems provide a viable approach to link the spatial-temporal flexibility with subjective wellbeing and potentially other socio-economic outcomes, notably labour productivity. ‘Back-to-work’ policies should provide wider support for lifestyle adaptation and transitions.

A significant limitation of the study is the single measurement of ExWB. A more comprehensive measure of subjective wellbeing would be ideal. This could potentially be achieved through implementing a supplemental questionnaire on long-term mental wellbeing following the original time-use survey. Standardised TUS data is collected in more than 30 countries. If more countries incorporate an ordinal ExWB variable and an additional questionnaire on long-term mental wellbeing, a large-scale international comparison could be conducted, which may shed light on the changing future of work from a public health perspective. Furthermore, the data from the UKTUS are population-representative but nevertheless cross-sectional. Higher quality longitudinal data, such as a cohort study would enable modelling analysis of causal relationship between lifestyles and ExWB.

### Funding and competing interest

This study received no external funding. The authors have declared that no competing interests exist.

### Research ethics approval

Ethics approval for this study was not required. All data involved is openly available to the public before the initiation of the study. The public data is hosted by UK Data Service, and can be accessed via the DOIs below: http://doi.org/10.5255/UKDA-SN-8128-1 http://doi.org/10.5255/UKDA-SN-8741-3

### Code availability

All STATA codes used for our data analysis are made publicly available on the project GitHub repository, and can be accessed via the URL below:

https://github.com/jerchenn/TimeUseWellbeing

## Data Availability

All data produced are available online at UK Data Service safeguarded depository.

https://beta.ukdataservice.ac.uk/datacatalogue/studies/study?id=8128

## APPENDICES

**Appendix 1:**
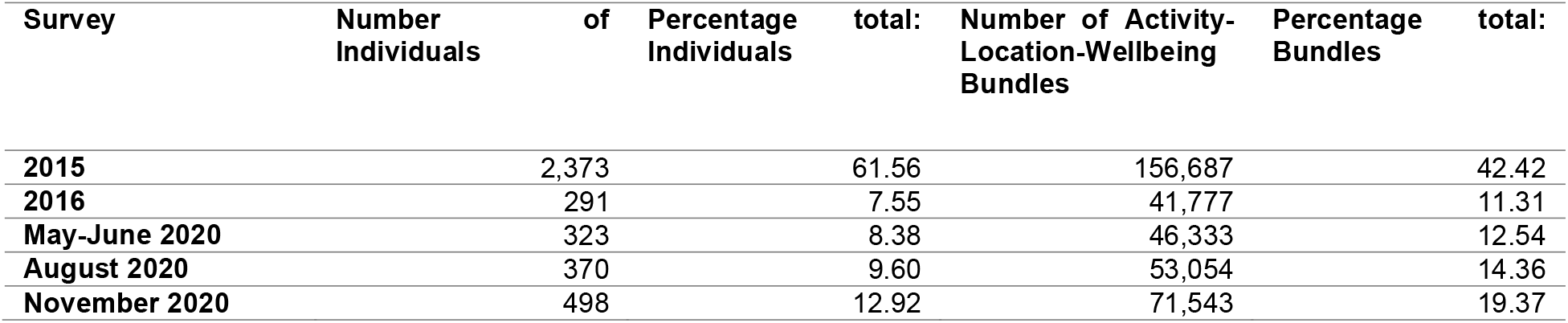
Sample distribution amongst survey waves

| Survey | Number<br>Individuals | of | Percentage<br>Individuals | total: | Number of Activity-Location-Wellbeing<br>Bundles | Percentage<br>Bundles | total: |
| --- | --- | --- | --- | --- | --- | --- | --- |
| 2015 | 2,373 |  | 61.56 |  | 156,687 |  | 42.42 |
| 2016 | 291 |  | 7.55 |  | 41,777 |  | 11.31 |
| May-June 2020 | 323 |  | 8.38 |  | 46,333 |  | 12.54 |
| August 2020 | 370 |  | 9.60 |  | 53,054 |  | 14.36 |
| November 2020 | 498 |  | 12.92 |  | 71,543 |  | 19.37 |

**Appendix 2:**
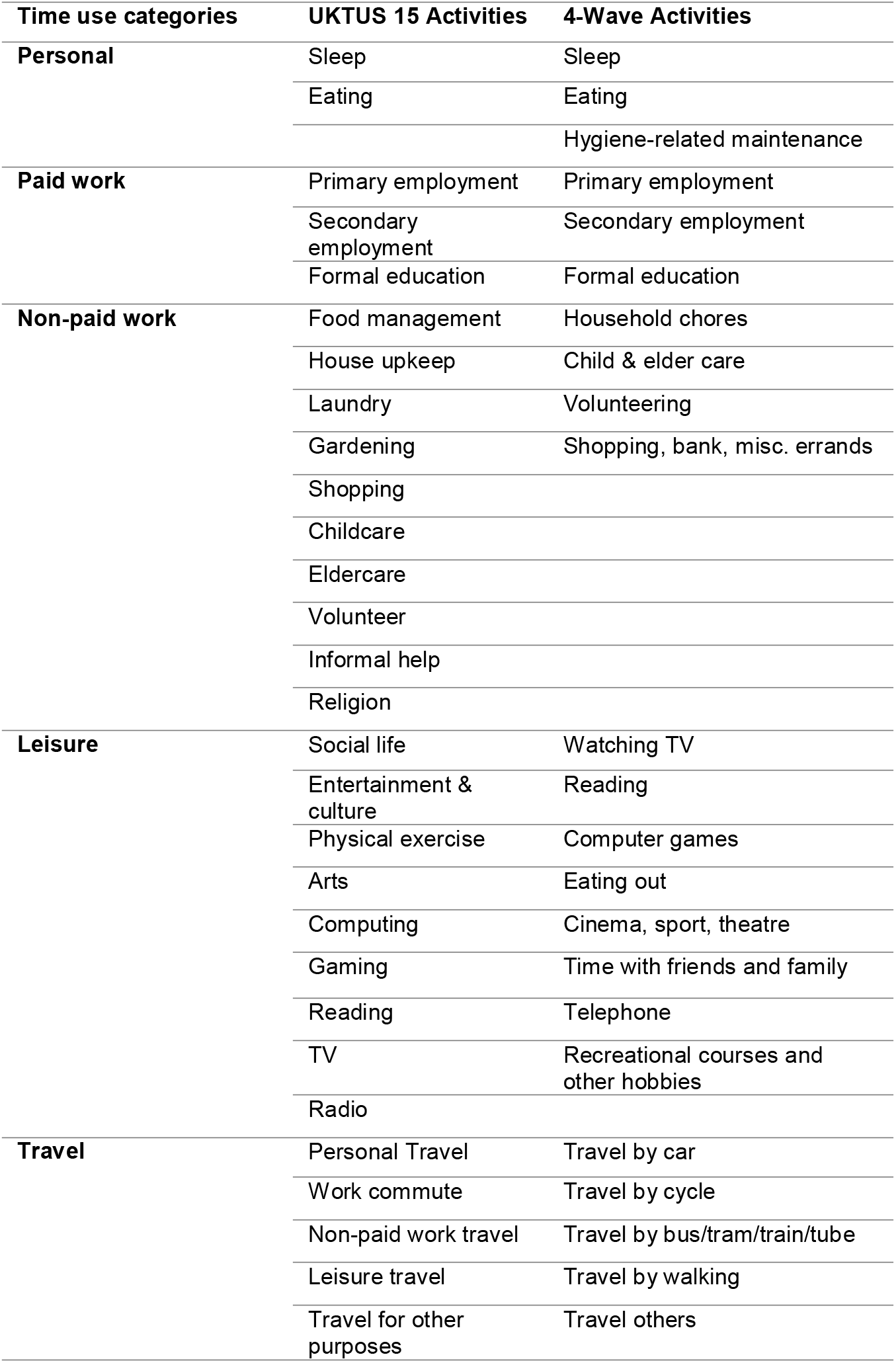
Aggregating activities into time use categories

**Appendix 3:**
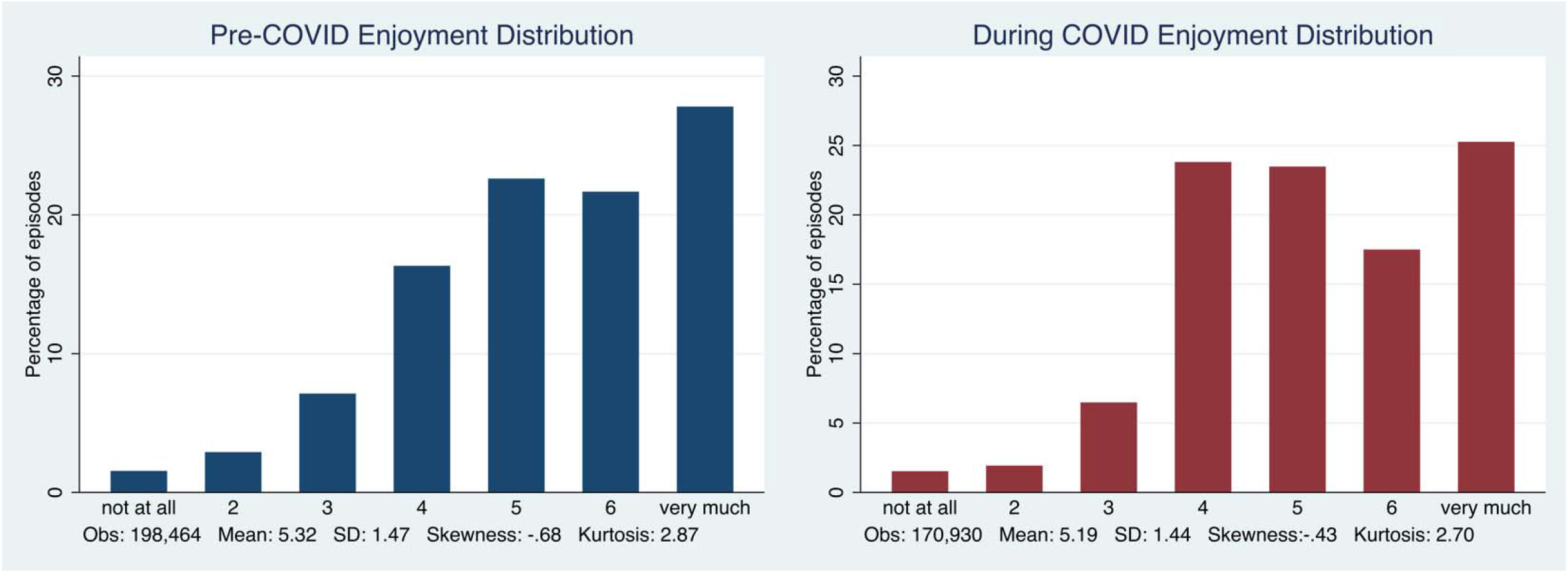
ExWB distribution

**Appendix 4:**
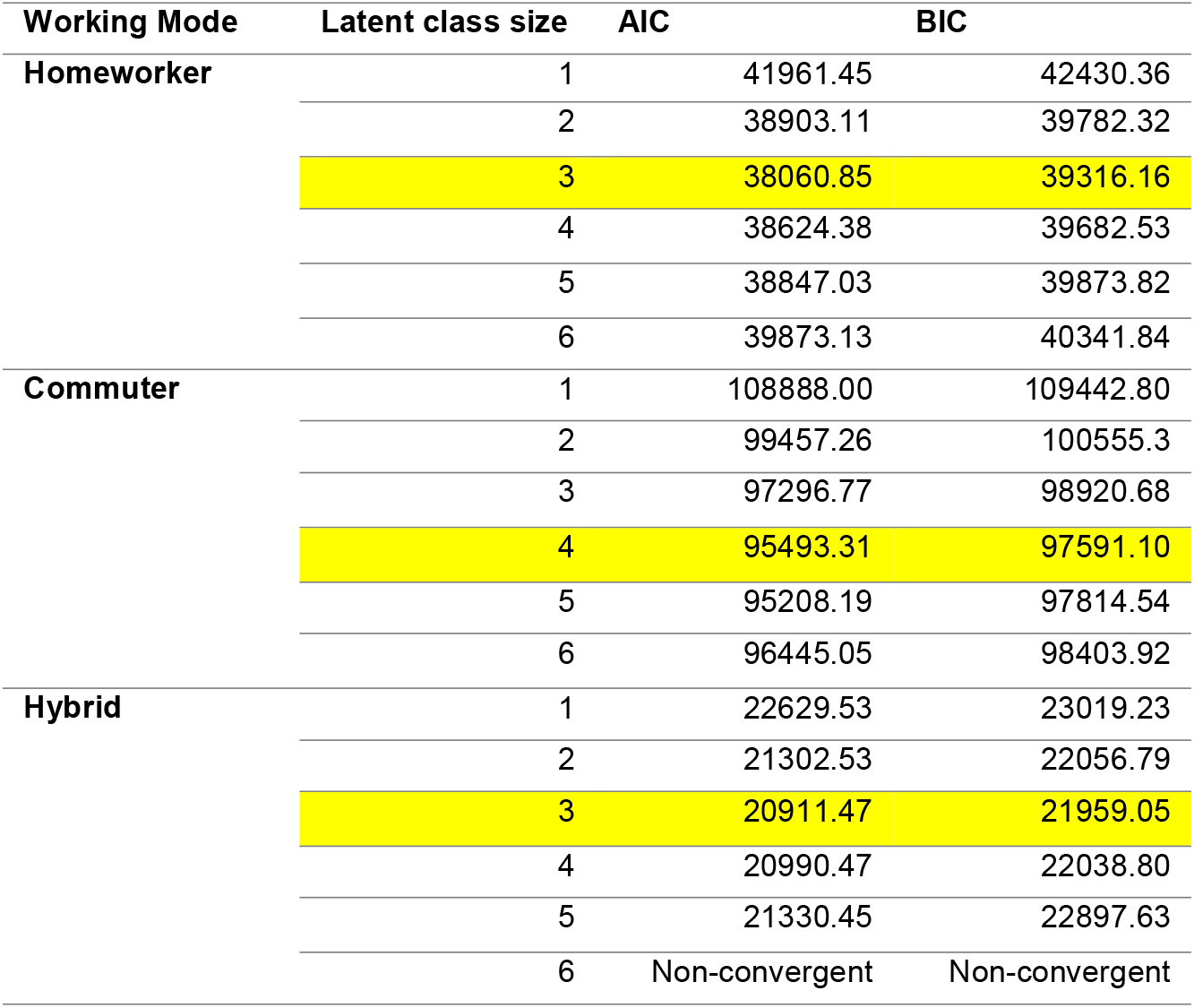
LCA model fit – information criterion

**Appendix 5:**
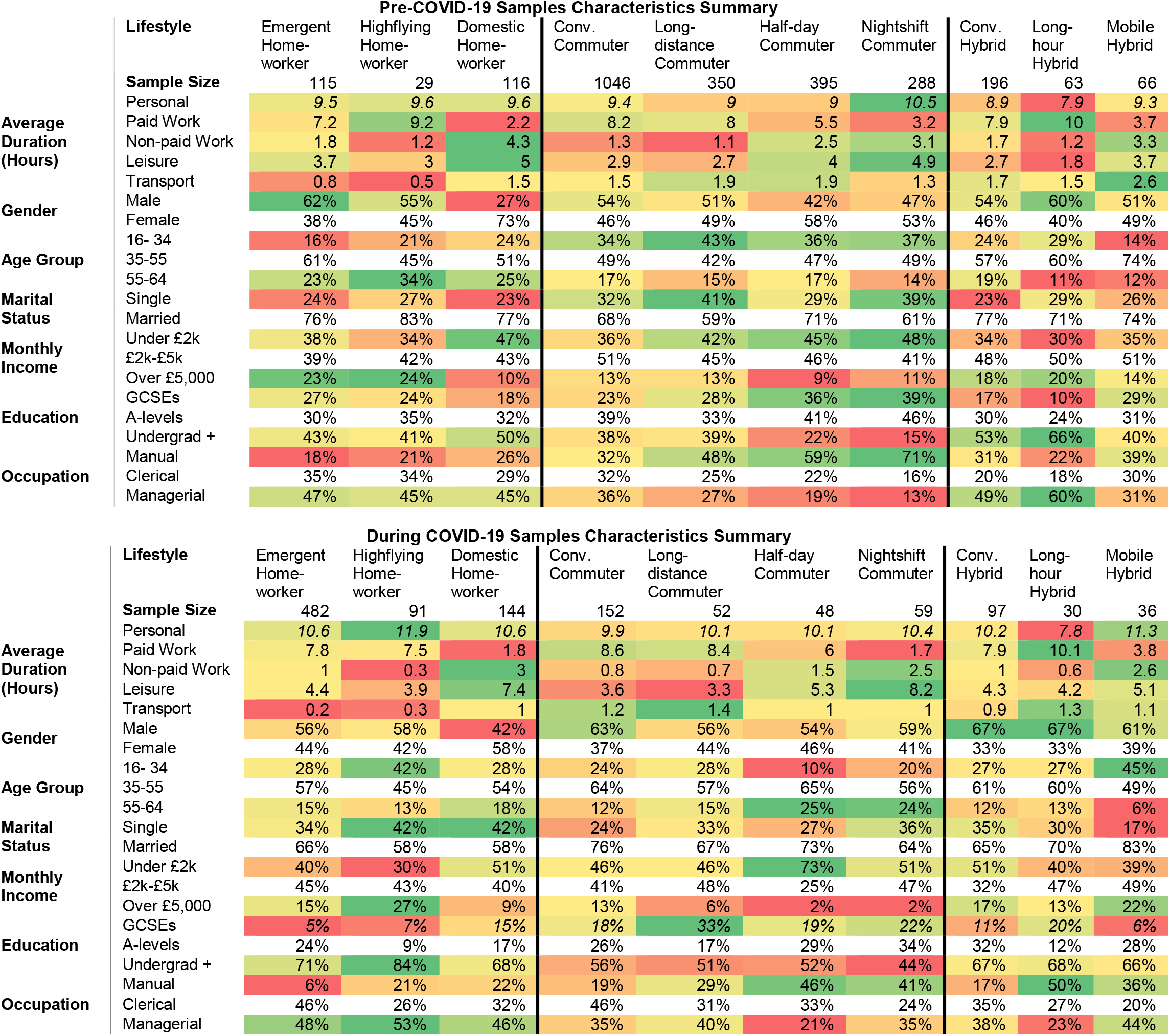
Sample descriptive statistics

