## Appendices for "Remote working and experiential wellbeing: A latent lifestyle perspective using UK Time Use Survey before and during COVID-19"

*Appendix 1: Sample distribution amongst survey waves*

| Survey | Number of Individuals | Percentage total: Individuals | Number of Activity-Location-Wellbeing Bundles | Percentage total: Bundles |
| --- | --- | --- | --- | --- |
| 2015 | 2,373 | 61.56 | 156,687 | 42.42 |
| 2016 | 291 | 7.55 | 41,777 | 11.31 |
| May-June 2020 | 323 | 8.38 | 46,333 | 12.54 |
| August 2020 | 370 | 9.60 | 53,054 | 14.36 |
| November 2020 | 498 | 12.92 | 71,543 | 19.37 |

*Appendix 2: Aggregating activities into time use categories*

| Time use categories | UKTUS 15 Activities | 4-Wave Activities |
| --- | --- | --- |
| Personal | Sleep | Sleep |
|  | Eating | Eating |
|  |  | Hygiene-related maintenance |
| Paid work | Primary employment | Primary employment |
|  | Secondary employment | Secondary employment |
|  | Formal education | Formal education |
| Non-paid work | Food management | Household chores |
|  | House upkeep | Child & elder care |
|  | Laundry | Volunteering |
|  | Gardening | Shopping, bank, misc. errands |
|  | Shopping |  |
|  | Childcare |  |
|  | Eldercare |  |
|  | Volunteer |  |
|  | Informal help |  |
|  | Religion |  |
| Leisure | Social life | Watching TV |
|  | Entertainment & culture | Reading |
|  | Physical exercise | Computer games |
|  | Arts | Eating out |
|  | Computing | Cinema, sport, theatre |
|  | Gaming | Time with friends and family |
|  | Reading | Telephone |
|  | TV | Recreational courses and other hobbies |
|  | Radio |  |
| Travel | Personal Travel | Travel by car |
|  | Work commute | Travel by cycle |
|  | Non-paid work travel | Travel by bus/tram/train/tube |
|  | Leisure travel | Travel by walking |
|  | Travel for other purposes | Travel others |

*Appendix 3: ExWB distribution*

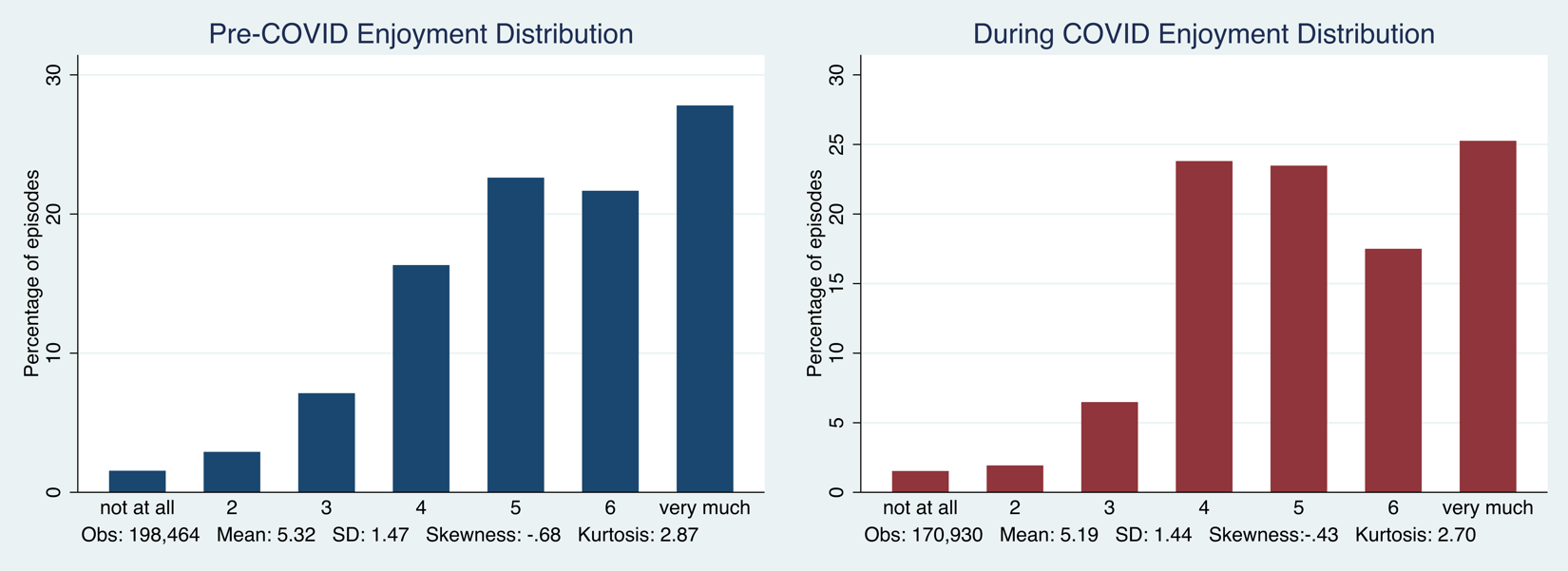

*Appendix 4: LCA model fit – information criterion*

| Working Mode | Latent class size | AIC | BIC |
| --- | --- | --- | --- |
| Homeworker | 1 | 41961.45 | 42430.36 |
|  | 2 | 38903.11 | 39782.32 |
|  | 3 | 38060.85 | 39316.16 |
|  | 4 | 38624.38 | 39682.53 |
|  | 5 | 38847.03 | 39873.82 |
|  | 6 | 39873.13 | 40341.84 |
| Commuter | 1 | 108888.00 | 109442.80 |
|  | 2 | 99457.26 | 100555.3 |
|  | 3 | 97296.77 | 98920.68 |
|  | 4 | 95493.31 | 97591.10 |
|  | 5 | 95208.19 | 97814.54 |
|  | 6 | 96445.05 | 98403.92 |
| Hybrid | 1 | 22629.53 | 23019.23 |
|  | 2 | 21302.53 | 22056.79 |
|  | 3 | 20911.47 | 21959.05 |
|  | 4 | 20990.47 | 22038.80 |
|  | 5 | 21330.45 | 22897.63 |
|  | 6 | Non-convergent | Non-convergent |

*Appendix 5: Sample descriptive statistics*

| **Pre-COVID-19 Samples Characteristics Summary** | | | | | | | | | | | |
| --- | --- | --- | --- | --- | --- | --- | --- | --- | --- | --- | --- |
|  | **Lifestyle** | Emergent Home-  worker | Highflying Home-  worker | Domestic Home-worker | Conv. Commuter | Long-distance Commuter | Half-day Commuter | Nightshift  Commuter | Conv. Hybrid | Long-hour Hybrid | Mobile Hybrid |
|  | **Sample Size** | 115 | 29 | 116 | 1046 | 350 | 395 | 288 | 196 | 63 | 66 |
| **Average Duration**  **(Hours)** | Personal | *9.5* | *9.6* | *9.6* | *9.4* | *9* | *9* | *10.5* | *8.9* | *7.9* | *9.3* |
|  | Paid Work | 7.2 | 9.2 | 2.2 | 8.2 | 8 | 5.5 | 3.2 | 7.9 | 10 | 3.7 |
|  | Non-paid Work | 1.8 | 1.2 | 4.3 | 1.3 | 1.1 | 2.5 | 3.1 | 1.7 | 1.2 | 3.3 |
|  | Leisure | 3.7 | 3 | 5 | 2.9 | 2.7 | 4 | 4.9 | 2.7 | 1.8 | 3.7 |
|  | Transport | 0.8 | 0.5 | 1.5 | 1.5 | 1.9 | 1.9 | 1.3 | 1.7 | 1.5 | 2.6 |
| **Gender** | Male | 62% | 55% | 27% | 54% | 51% | 42% | 47% | 54% | 60% | 51% |
|  | Female | 38% | 45% | 73% | 46% | 49% | 58% | 53% | 46% | 40% | 49% |
| **Age Group** | 16- 34 | 16% | 21% | 24% | 34% | 43% | 36% | 37% | 24% | 29% | 14% |
|  | 35-55 | 61% | 45% | 51% | 49% | 42% | 47% | 49% | 57% | 60% | 74% |
|  | 55-64 | 23% | 34% | 25% | 17% | 15% | 17% | 14% | 19% | 11% | 12% |
| **Marital Status** | Single | 24% | 27% | 23% | 32% | 41% | 29% | 39% | 23% | 29% | 26% |
|  | Married | 76% | 83% | 77% | 68% | 59% | 71% | 61% | 77% | 71% | 74% |
| **Monthly Income** | Under £2k | 38% | 34% | 47% | 36% | 42% | 45% | 48% | 34% | 30% | 35% |
|  | £2k-£5k | 39% | 42% | 43% | 51% | 45% | 46% | 41% | 48% | 50% | 51% |
|  | Over £5,000 | 23% | 24% | 10% | 13% | 13% | 9% | 11% | 18% | 20% | 14% |
| **Education** | GCSEs | 27% | 24% | 18% | 23% | 28% | 36% | 39% | 17% | 10% | 29% |
|  | A-levels | 30% | 35% | 32% | 39% | 33% | 41% | 46% | 30% | 24% | 31% |
|  | Undergrad + | 43% | 41% | 50% | 38% | 39% | 22% | 15% | 53% | 66% | 40% |
| **Occupation** | Manual | 18% | 21% | 26% | 32% | 48% | 59% | 71% | 31% | 22% | 39% |
|  | Clerical | 35% | 34% | 29% | 32% | 25% | 22% | 16% | 20% | 18% | 30% |
|  | Managerial | 47% | 45% | 45% | 36% | 27% | 19% | 13% | 49% | 60% | 31% |

| **During COVID-19 Samples Characteristics Summary** | | | | | | | | | | | |
| --- | --- | --- | --- | --- | --- | --- | --- | --- | --- | --- | --- |
|  | **Lifestyle** | Emergent Home-  worker | Highflying Home-  worker | Domestic Home-worker | Conv.  Commuter | Long-distance Commuter | Half-day Commuter | Nightshift  Commuter | Conv. Hybrid | Long-hour Hybrid | Mobile Hybrid |
|  | **Sample Size** | 482 | 91 | 144 | 152 | 52 | 48 | 59 | 97 | 30 | 36 |
| **Average Duration**  **(Hours)** | Personal | *10.6* | *11.9* | *10.6* | *9.9* | *10.1* | *10.1* | *10.4* | *10.2* | *7.8* | *11.3* |
|  | Paid Work | 7.8 | 7.5 | 1.8 | 8.6 | 8.4 | 6 | 1.7 | 7.9 | 10.1 | 3.8 |
|  | Non-paid Work | 1 | 0.3 | 3 | 0.8 | 0.7 | 1.5 | 2.5 | 1 | 0.6 | 2.6 |
|  | Leisure | 4.4 | 3.9 | 7.4 | 3.6 | 3.3 | 5.3 | 8.2 | 4.3 | 4.2 | 5.1 |
|  | Transport | 0.2 | 0.3 | 1 | 1.2 | 1.4 | 1 | 1 | 0.9 | 1.3 | 1.1 |
| **Gender** | Male | 56% | 58% | 42% | 63% | 56% | 54% | 59% | 67% | 67% | 61% |
|  | Female | 44% | 42% | 58% | 37% | 44% | 46% | 41% | 33% | 33% | 39% |
| **Age Group** | 16- 34 | 28% | 42% | 28% | 24% | 28% | 10% | 20% | 27% | 27% | 45% |
|  | 35-55 | 57% | 45% | 54% | 64% | 57% | 65% | 56% | 61% | 60% | 49% |
|  | 55-64 | 15% | 13% | 18% | 12% | 15% | 25% | 24% | 12% | 13% | 6% |
| **Marital Status** | Single | 34% | 42% | 42% | 24% | 33% | 27% | 36% | 35% | 30% | 17% |
|  | Married | 66% | 58% | 58% | 76% | 67% | 73% | 64% | 65% | 70% | 83% |
| **Monthly Income** | Under £2k | 40% | 30% | 51% | 46% | 46% | 73% | 51% | 51% | 40% | 39% |
|  | £2k-£5k | 45% | 43% | 40% | 41% | 48% | 25% | 47% | 32% | 47% | 49% |
|  | Over £5,000 | 15% | 27% | 9% | 13% | 6% | 2% | 2% | 17% | 13% | 22% |
| **Education** | GCSEs | *5%* | *7%* | *15%* | *18%* | *33%* | *19%* | *22%* | *11%* | *20%* | *6%* |
|  | A-levels | 24% | 9% | 17% | 26% | 17% | 29% | 34% | 32% | 12% | 28% |
|  | Undergrad + | 71% | 84% | 68% | 56% | 51% | 52% | 44% | 67% | 68% | 66% |
| **Occupation** | Manual | 6% | 21% | 22% | 19% | 29% | 46% | 41% | 17% | 50% | 36% |
|  | Clerical | 46% | 26% | 32% | 46% | 31% | 33% | 24% | 35% | 27% | 20% |
|  | Managerial | 48% | 53% | 46% | 35% | 40% | 21% | 35% | 38% | 23% | 44% |
